# PPI-Refractory GERD in Systemic Sclerosis Is Driven by Distinct Esophageal and Gastric Motility Abnormalities

**DOI:** 10.64898/2026.04.13.26350585

**Authors:** Luis G Alcala-Gonzalez, Alfredo Guillen-del-Castillo, Francisco Alejandro Felix Tellez, Ariadna Aguilar, Claudia Barber-Caselles, Carolina Malagelada, Laura Polo Figueras, Laura Triginer, Claudia Codina-Clavaguera, Michael Hughes, Carmen Pilar Simeón-Aznar, Jordi Serra, Zsuzsanna H. McMahan

## Abstract

**Background:** Gastroesophageal reflux disease (GERD) is highly prevalent in systemic sclerosis (SSc) and frequently persists despite proton pump inhibitor (PPI) therapy. However, the mechanisms underlying PPI-refractory GERD in SSc remain incompletely understood.

**Methods:** We conducted a singlel7lcentre, retrospective study of adults with SSc who underwent ambulatory pH-multichannel intraluminal impedance (pH/MII) monitoring while receiving twicel7ldaily PPI therapy (2021–2025). Esophageal motility (highl7lresolution manometry, HREM) and gastric emptying scintigraphy were integrated to examine associations between gastro-esophageal dysmotility and reflux phenotypes.

**Results:** Thirty patients were included, of whom 67% had PPI-refractory reflux symptoms and 33% were undergoing pre–lung transplantation evaluation. Refractory GERD was present in 29/30 patients (97%) based on Lyon 2.0 classification, with conclusive evidence in 53% and borderline evidence in 43%. Esophageal dysmotility was identified in 80%, most commonly absent contractility (67%), and was associated with impaired reflux clearance, reflected by longer acid clearance times (2.20 [1.15–3.75] vs 1.15 [0.43–1.90] min) and prolonged reflux episode duration (16.60 [4.38–40.63] vs 1.95 [0.53–20.43] min). Gastric dysmotility was identified in 60.7% and was associated with an increased reflux episode burden (51.00 [30.00–81.50] vs 25.00 [21.00–54.00] episodes/24h).

**Conclusions:** PPIl7lrefractory GERD is nearly universal in this SSc cohort and reflects heterogeneous, quantifiable abnormalities across the foregut, including impaired esophageal clearance and increased reflux burden related to gastric retention. These findings support integrated physiologic evaluation to define reflux mechanisms, inform risk stratification (including lung transplantation), and guide targeted, mechanism-based therapies beyond acid suppression.

## INTRODUCTION

Systemic sclerosis (SSc) is a multisystem rheumatological disease characterized by vasculopathy, fibrosis, and immune dysregulation, with gastrointestinal (GI) involvement affecting up to 90% of patients over the disease course^1^. Among all extracutaneous manifestations, GI dysmotility is both the most common and one of the most debilitating, producing clinically significant complications across the GI tract, including gastro-esophageal reflux disease (GERD), gastroparesis, small intestinal bacterial overgrowth, and intestinal pseudo-obstruction^2,3^. Objective assessment of GI dysmotility using highl7lresolution manometry (HREM), radiologic imaging, and transit studies has become central to understanding SScl7lrelated GI disease, yet major questions remain regarding how specific motility disturbances contribute to key clinical outcomes^4,5^.

GERD is one of the most frequent and clinically impactful GI manifestations in SSc^6,7^. Excess esophageal exposure to gastric refluxate is the primary driver of mucosal injury and contributes to significant downstream complications such as erosive esophagitis, Barrett’s esophagus, adenocarcinoma, and extraesophageal reflux burden involving the larynx and airways^8,9^. Of particular importance in SSc, microaspiration may accelerate interstitial lung disease (ILD) progression and worsen postl7llung transplant outcomes, making effective reflux control a critical component of disease management^10^. Although proton pump inhibitors (PPIs) remain the cornerstone of therapy and prevent acidl7lmediated injury in the general population, a substantial proportion of SSc patients continue to exhibit objective evidence of reflux pathology despite optimized PPI therapy^7,11,12^. Furthermore, concerns have been raised around the safety of long-term PPI exposure in patients with SSc (mainly extrapolated from the general population)^13,14^. Therefore, treatmentl7lrefractory GERD represents a clinically meaningful and understudied challenge.

The mechanisms underlying PPIl7lrefractory GERD in SSc are incompletely understood. Prior work has largely focused on esophageal motor dysfunction, particularly absent contractility, which contributes to impaired reflux clearance and greater esophageal acid exposure^15,16^. However, the relationship between esophageal dysmotility and GERD severity in SSc is inconsistent, and esophageal findings alone do not fully explain persistent symptoms or mucosal injury. Emerging data suggest that gastric and small bowel dysmotility may independently contribute to increased reflux burden, nonl7lacid reflux exposure, and the persistence of symptoms despite adequate acid suppression^11,17^. Yet, these mechanisms have rarely been studied together, and no prior work has integrated comprehensive gastro-esophageal motility testing with pH-multichannel intraluminal impedance (pH/MII) monitoring performed during PPI therapy, the condition under which refractory symptoms actually occur.

This lack of integrated physiologic evaluation represents a major gap in the extant literature and has practical implications for real-world management^18^. Current GERD diagnostic algorithms, developed primarily in nonl7lSSc populations, recommend offl7lPPI pH testing to confirm GERD before considering refractory disease^19,20^. However, discontinuing PPIs is often poorly tolerated in SSc due to the presence of severe reflux, and the underlying mechanisms of reflux in this population differ markedly from typical GERD^13^. As a result, objective evaluation strategies validated in nonl7lSSc populations may be inadequate or inappropriate. There is an urgent need to define how gastro-esophageal physiological disturbances manifest on PPI therapy, when treatment decisions must be made.

Thus, the goal of this study was to address this critical knowledge gap by comprehensively characterizing reflux physiology using pH/MII monitoring performed during ongoing PPI therapy and by integrating these findings with objective measures of esophageal and gastric motility. Specifically, we aimed to (1) define the prevalence and characteristics of PPIl7lrefractory GERD in SSc under reall7lworld treatment conditions, and (2) elucidate the relative contributions of esophageal dysmotility, gastric emptying abnormalities, and EGJ barrier dysfunction to persistent reflux burden. By linking these physiologic domains, our study should provide new mechanistic insight into refractory GERD in SSc and establish a clinically actionable framework for mechanisml7lbased evaluation and management.

## METHODS

### Study design and setting

We performed a single-centre, retrospective, cross-sectional study in patients from the Scleroderma Unit cohort at the University Hospital Vall d’Hebron who met the ACR/EULAR 2013 criteria^21^ and/or LeRoy classification criteria^22^. We included all patients who underwent pH/MII monitoring on twice-daily PPI therapy between 2021 and 2025. Patients were evaluated for ongoing GERD in one of two scenarios: (a) persistent bothersome reflux symptoms, defined as ongoing esophageal or extra-esophageal symptoms despite twice-daily PPI therapy; or (b) pre–lung transplant assessment, including patients with typical reflux symptoms undergoing evaluation irrespective of symptom refractoriness.

Patients with prior upper gastrointestinal surgical or endoscopic interventions (e.g., fundoplication, stricture dilation, endoscopic anti-reflux procedures) were excluded. Clinical, demographic, immunological characteristics and results from GI motility tests were retrieved from a prospectively collected database, as defined in previous reports^4,5,23^. All patients provided written informed consent for the collection and analysis of their clinical data [PR(AG)312/2021].

### 24-hour ambulatory pH–impedance procedure

Impedance measurement is based on the principle that the various contents of the esophageal lumen (air, liquid, solids, mucus) have different electrical conductivities, measured as impedance (resistance Ω) between pairs of consecutive electrodes on a multi-channel catheter. Ambulatory pH/MII study was performed in all patients using a Zephyr multi-channel intraluminal impedance ambulatory system (ZepHr® Impedance/pH, Diversatek Healthcare, Boulder, CO, USA), consisting of six impedance channels and a distal pH sensor. The catheter was positioned with the pH electrode 5 cm above the upper border of the lower esophageal sphincter (LES), as previously determined by high-resolution esophageal manometry (HREM). During the study, patients kept a standardized diary recording meal times, body position (supine/upright), and the occurrence of symptoms (heartburn, regurgitation, chest pain, cough, and/or throat symptoms). Patients returned the equipment and diary the following day. Meal periods were excluded from the analysis.

Recordings were visually analyzed using dedicated software (Zvu® Analysis, version 3.2.0; Diversatek Healthcare, Boulder, CO, USA). All tracings were independently reviewed by two experienced gastroenterologist investigators (FT and LGAG). Reflux episodes were identified according to the Wingate consensus criteria^24^. Acid exposure time was calculated as the percentage of time with esophageal pH <4 in the distal esophagus during the recording period. Additional parameters included the longest acid exposure time (minutes), median bolus clearance time (seconds), and mean nocturnal baseline impedance (MNBI), calculated as the mean of three measurements obtained during nocturnal supine periods after sleep onset^25^. Reflux–symptom association was considered positive when either the symptom index (SI ≥50%) or the symptom association probability (SAP ≥95%) was abnormal^26^.

### Gastrointestinal involvement evaluation at inclusion

At our center, all patients with SSc systematically undergo esophageal and gastric motility testing as part of a standardized GI assessment protocol. Esophageal motility was evaluated using HREM. All tracings were reviewed and interpreted according to the Chicago Classification 4.0^27^ by two experienced gastroenterologist investigators (FT and LGAG). The integrity of the esophagogastric junction (EGJ) anti-reflux barrier was evaluated using the Milan score metrics^28^, which reflect its functional competence. EGJ barrier function was quantified by measuring the EGJ contractile integral (EGJ-CI) and by assessing its dynamic competence during the supine-to-upright transition maneuver^29^. These parameters were integrated with esophageal motility to calculate the Milan score^28^, a screening tool to stratify the risk and the severity of GERD based on HREM. Gastric motor function was assessed using gastric emptying scintigraphy with a standardized solid meal. Gastric emptying was quantified as the proportion of meal retention at 4 hours and categorized according to established reference thresholds from our lab^4,30^. Detailed descriptions of the motility protocols and analysis of gastro-esophageal motility tests are provided in the Supplementary Methods.

### Outcomes definitions

The primary outcome was the prevalence of PPI-refractory GERD, defined using an operational framework adapted from the Lyon 2.0 as either conclusive evidence (acid exposure time >4% and/or >80 reflux episodes per day) or borderline evidence (acid exposure time 1–4%, 40–80 reflux episodes per day, or MNBI <2500 Ω) suggestive of ongoing reflux despite PPI therapy^19^. GERD was considered controlled when pH/MII findings met all criteria against ongoing reflux while on PPI therapy (acid exposure time <1%, <40 reflux episodes per day, and MNBI >2500 Ω). Secondary outcomes were exploratory and included the prevalence of gastro-esophageal dysmotility and its association with pH/MII parameters. To allow comparison across different PPIs, doses were standardized using omeprazole-equivalent (OE) dosing according to previously published conversion factors^31^. Daily PPI exposure was then categorized as low dose (≤20 mg OE), full dose (21–40 mg OE), or high dose (>40 mg OE).

### Statistical Analysis

All statistical analyses were performed using JASP Version 0.18.3 (JASP TEAM, Amsterdam, The Netherlands). The normality of continuous variables was assessed using the Shapiro–Wilk test. Continuous variables are reported as mean ± SD or median (IQR); categorical variables as n (%). Group comparisons used t-test or Mann–Whitney U for continuous variables and χ²/Fisher’s exact for categorical variables. Correlations used Spearman’s ρ. A two-sided P-value <0.05 was considered statistically significant.

## RESULTS

### Patient characteristics

Of the 40 patients with SSc who underwent pH/MII monitoring while on PPI therapy, 10 were excluded: six were not receiving twicel7ldaily PPI at the time of testing, three had prior gastroesophageal or relevant thoracic surgery (two fundoplications and one lung transplantation), and one was receiving concomitant GLPl7l1 receptor agonist therapy. The final analytic cohort comprised 30 patients (**Table 1**). All participants reported esophageal symptoms at baseline, most commonly regurgitation (n=22) and heartburn (n=20), with fewer reporting dysphagia (n=7) or chest pain (n=3). pH/MII monitoring was performed to evaluate persistent reflux symptoms in 20 patients (66.7%), while the remaining 10 (33.3%) underwent testing as part of pre–lung transplantation assessment despite lack of ongoing symptoms. Omeprazole was the most commonly prescribed PPI (n=13, 43.3%). After conversion to OE dosing, the median daily PPI exposure was 40 mg OE (IQR 27–64; range 18–72). Half of the cohort (n=15, 50.0%) received fulll7ldose (21–40 mg OE), 12 patients (40.0%) were treated with highl7ldose (>40 mg OE), and three (10.0%) received lowl7ldose therapy (≤20 mg OE).

**Table 1:**
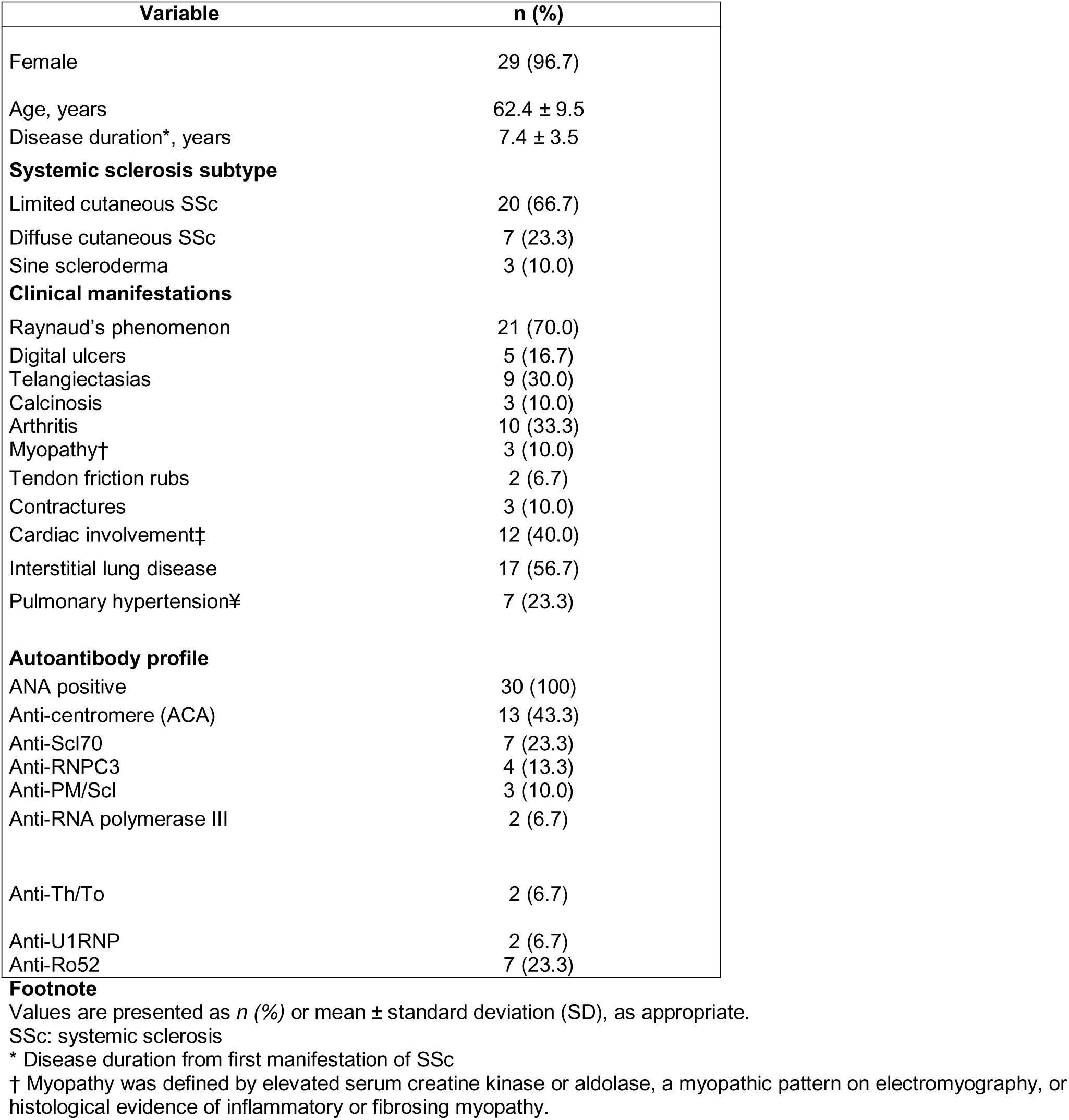
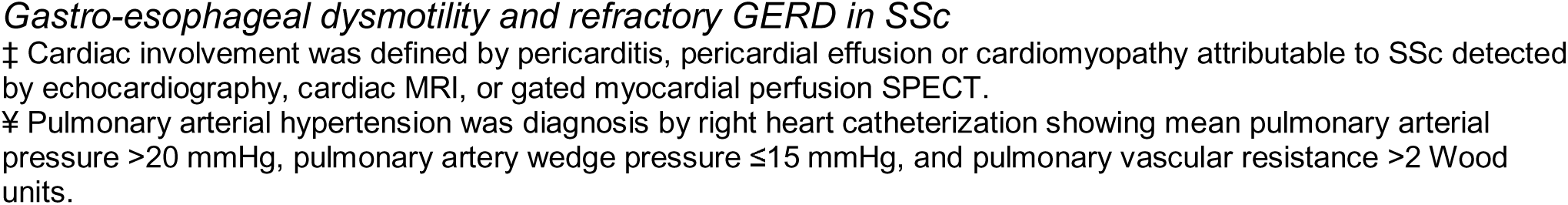
Demographic and clinical characteristics of included patients.

### Evaluation of ongoing GERD despite PPI therapy

The distribution of pH/MII outcomes is shown in Figure 1, with panel A depicting the prevalence of abnormalities across individual metrics and panel B showing patient-level transitions and final GERD classification using an alluvial plot. Nearly all patients demonstrated objective evidence of ongoing reflux despite PPI therapy: 29/30 (97%) met criteria for refractory GERD, including 16 (53%) with conclusive and 13 (43%) with borderline abnormalities. Only one patient (3%) had findings consistent with adequate reflux control on twice-daily PPI therapy. Detailed pH/MII parameters metrics are presented in **Table 2**. Abnormal acid exposure was common, affecting 19 patients (63%), including 14 (47%) with conclusive acid exposure time >4% despite PPI therapy. The number of reflux episodes was also frequently elevated: 4 patients (13%) exhibited conclusive increases in total reflux episodes (>80 episodes/24 h), and 10 (33 %) had borderline results (>40 episodes/24 hrs).

**Figure 1:**
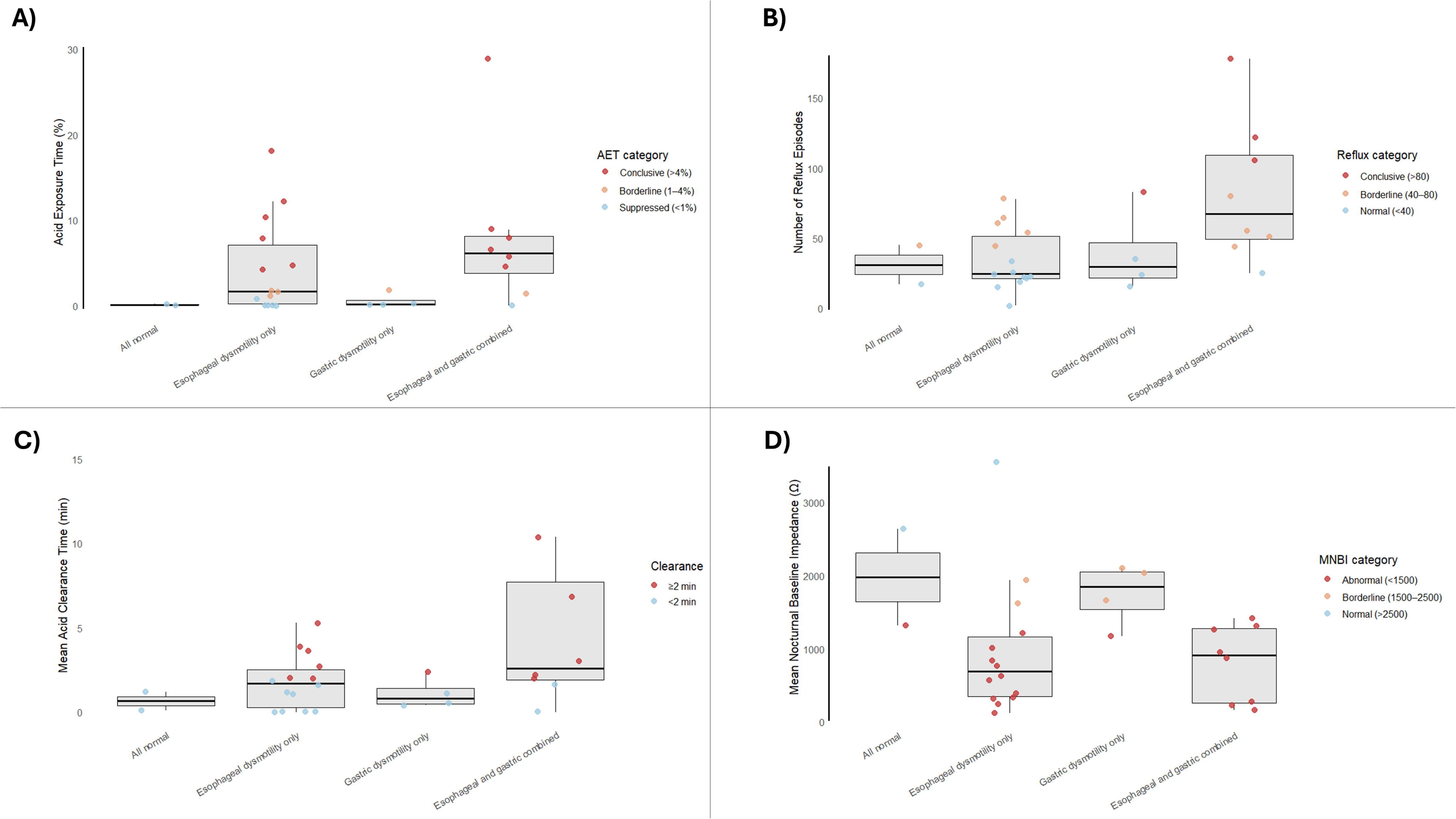
Prevalence of pH/MII abnormalities (A) and patient-level trajectories to final pH/MII classification (B). **Footnote:** Acid exposure time (AET) was categorized as suppressed (<1%), borderline (1–4%), or conclusive (>4%). Total reflux episodes were classified as normal (<40), borderline (40–80), or conclusive (>80). Mean nocturnal baseline impedance (MNBI) was categorized as normal (>2500 Ω), borderline (1500–2500 Ω), or abnormal (<1500 Ω), reflecting impaired mucosal integrity. Overall pH/MII status was classified as controlled, borderline, or conclusive findings according to Lyon 2.0 criteria. In panel B, stream widths represent the number of patients transitioning between categories across metrics.

**Table 2:** pH-impedance parameters results in the whole cohort.

| Key Parameters Measured | Normal values | Median (IQR) |
| --- | --- | --- |
| Total acid exposure time (%/24h) | Normal < 1% (On PPI) | 1.75 (0.13–7.90) |
| Upright acid exposure time (%/24h) |  | 2.65 (0.23–7.03) |
| Supine acid exposure time (%/24h) |  | 0.10 (0.00–6.90) |
| Acid exposure duration (min) | No established reference | 22.75 (1.65–113.20) |
| DeMeester score | Normal < 14.72 | 5.90 (1.10–34.80) |
| Total impedance reflux episodes (n/24h) | Normal < 40 episodes/24hrs | 35.00 (22.25–59.50) |
| Acid reflux episodes (n/24h) |  | 3.00 (1.25–11.25) |
| Weakly acidic reflux episodes (n/24h) |  | 23.00 (16.00–39.75) |
| Mean acid clearance time (min) |  | 1.80 (0.50–2.70) |
| Longest acid reflux episode (min) |  | 10.50 (0.83–30.08) |
| Mean nocturnal baseline impedance (MNBI, Ω) | Abnormal < 2500 Ω | 986.50 (392–1392) |

Values are presented as median (interquartile range, IQR) and mean ± standard deviation (SD). Total acid exposure time (AET) represents the percentage of time with esophageal pH <4 over 24 hours. Upright and supine AET correspond to acid exposure during upright and recumbent periods, respectively. The DeMeester score is a composite metric of esophageal acid exposure, with values <14.72 considered normal. Impedance reflux episodes represent the total number of reflux events detected by impedance monitoring over 24 hours; Acid reflux episodes are defined as reflux events with pH <4, while weakly acidic reflux episodes are defined as events with pH between 4 and 7. Mean acid clearance time reflects the duration required for esophageal pH to return above 4 following a reflux event. The longest acid reflux episode represents the maximum duration of a single reflux event during the recording period. Mean nocturnal baseline impedance reflects esophageal mucosal integrity and/or stasis.

Abnormal MNBI was the most prevalent abnormality, <1500 Ω in 23 patients (77%), with an additional 5 patients (17%) exhibiting borderline values (1500-2500 Ω). Of the 17 patients who reported symptoms during monitoring, 8 (47%) had a positive reflux–symptom association, reinforcing the clinical relevance of the physiologic abnormalities detected.

Exploratory analyses demonstrated no statistically significant differences in pH/MII results categories in relation to PPI dose categories (p=0.450), although patients on highl7ldose PPI (>40 mg OE) showed a numerically higher prevalence of both conclusive and borderline acid exposure. Ro52 antibody positivity was more frequent among patients with conclusive findings of ongoing reflux (85.7% vs. 14.3%; p=0.050).

### Associations Between Gastro-Esophageal Motility and Refractory GERD

All patients underwent objective motility testing at baseline. HREM was performed a median of 12 months before pH/MII testing (IQR 0–20), and gastric emptying scintigraphy was available for 28 patients, conducted a median of 9 months before pH/MII (IQR 3–15). Abnormalities in esophageal motility were common, identified in 24 of 30 patients (80%), most frequently absent contractility (n=20; 67%) followed by ineffective esophageal motility (IEM; n=4, 13%). A hiatal hernia >2 cm was present in 11 patients (37%). Measures of EGJ barrier function were frequently abnormal, with a reduced EGJ contractile integral (EGJl7lCI <23 mmHg·cm) observed in 14 patients (47%).

Among patients with available scintigraphy data, delayed gastric emptying was identified in 17 of 28 (61%), including mild delay (10-20% retention at 4 h; n=6, 21%), moderate delay (20-35%; n=6, 21%), and severe delay (>35%; n=5, 18%).

Figures 2 illustrates the distribution of gastro-esophageal motility abnormalities across categories of esophageal acid exposure time (AET; panel A) and reflux episode burden (panel B), highlighting distinct associations between reflux metrics and esophageal versus gastric dysfunction.

**Figure 2.**
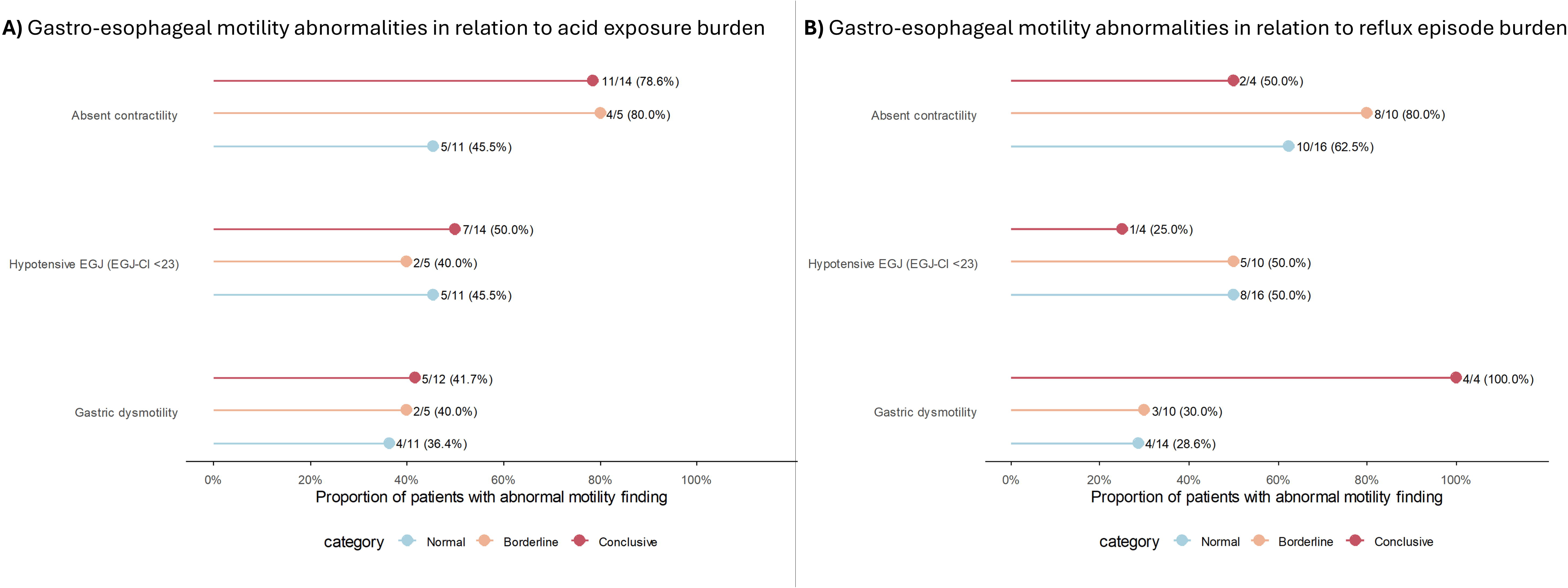
Gastro-esophageal motility abnormalities according to esophageal acid exposure (A) and reflux episode burden (B). Footnote: Proportions represent the percentage of patients within each category who exhibited the corresponding motility abnormality, with absolute numbers shown for each subgroup. Acid exposure and reflux episode burden categories were defined according to Lyon 2.0 criteria. Across both panels, motility abnormalities showed a non-uniform distribution across reflux severity strata. Elevated acid exposure was predominantly associated with absent esophageal contractility, whereas higher reflux episode burden was more frequently associated with gastric dysmotility, supporting distinct pathophysiologic contributions to reflux phenotypes.

**Table 3** summarizes pH-impedance metrics stratified by esophageal motility subtypes and gastric motor function. Across esophageal motility categories, a graded pattern was observed from normal motility to IEM and absent contractility, with progressively higher AET (0.15 [0.10–1.40] vs 4.55 [0.15–9.25] vs 4.40 [1.03–7.90]%), and prolonged acid clearance times (0.80 [0.43–1.48] vs 1.60 [0.90–2.00] vs 2.20 [1.15–3.75] min). In contrast, gastric dysmotility was primarily associated with increased reflux episode burden. Patients with delayed gastric emptying exhibited a greater total number of reflux episodes (51.00 [30.00–81.50] vs 25.00 [21.00–54.00] episodes/24 h), driven largely by weakly acidic reflux (34.00 [20.00–67.50] vs 21.00 [16.00–39.00] episodes/24 h), with less pronounced differences in AET.

**Table 3:**
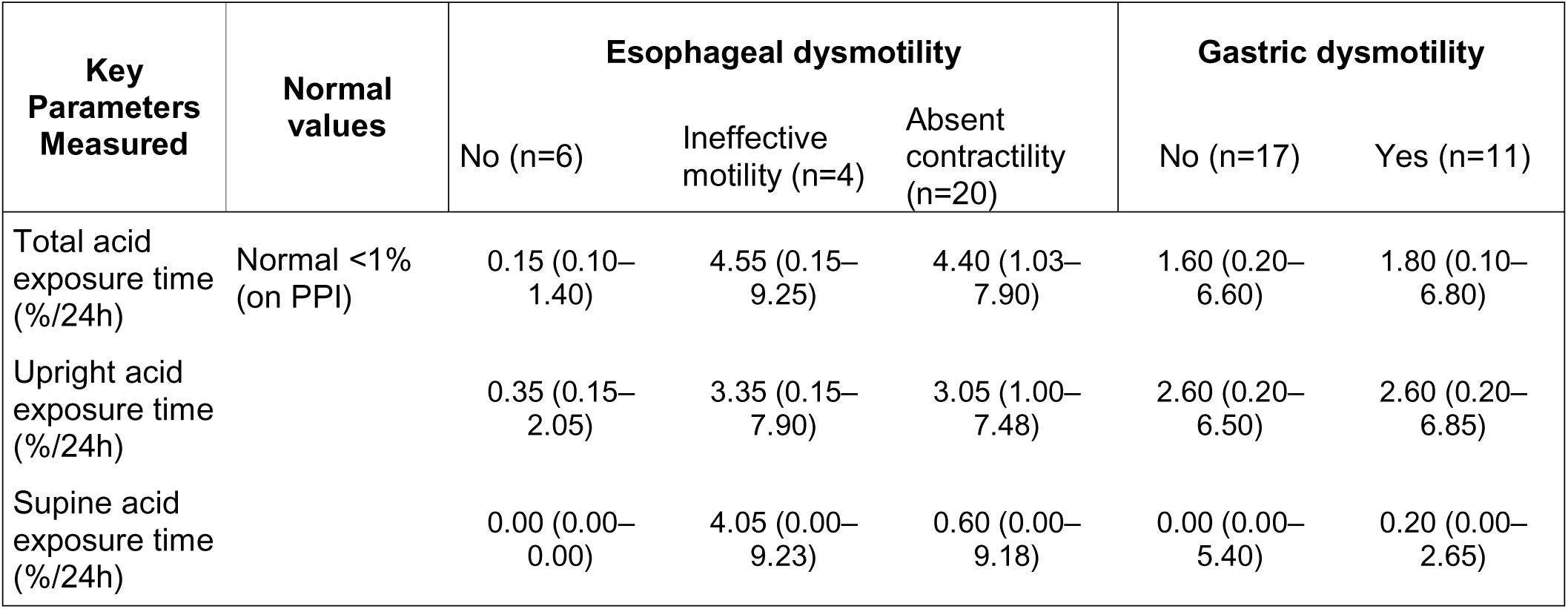
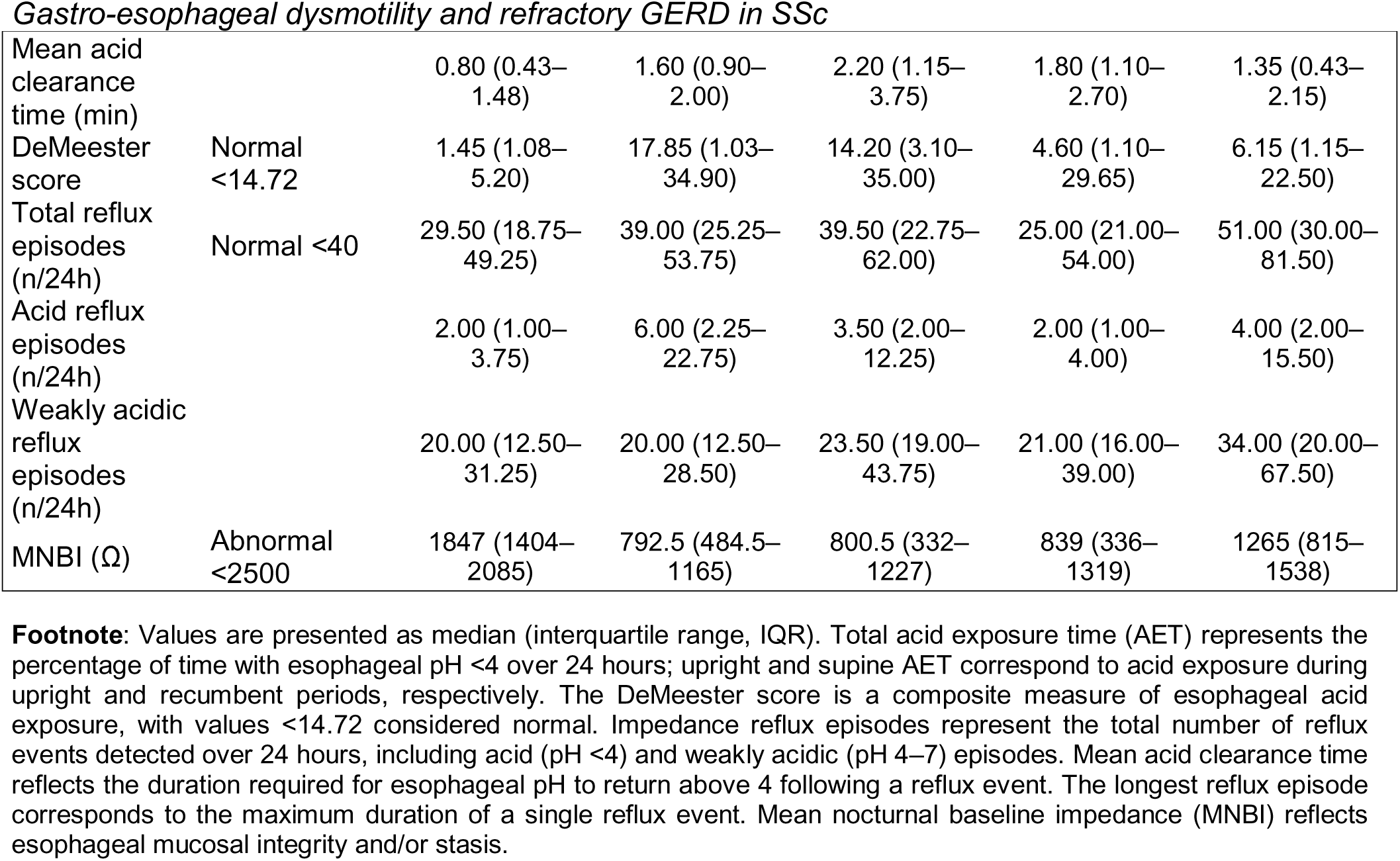
pH-impedance parameters according to esophageal and gastric motility findings.

| Key Parameters Measured | Normal values | Esophageal dysmotility |  |  | Gastric dysmotility |  |
| --- | --- | --- | --- | --- | --- | --- |
|  |  | No (n=6) | Ineffective motility (n=4) | Absent contractility (n=20) | No (n=17) | Yes (n=11) |
| Total acid exposure time (%/24h) | Normal <1% (on PPI) | 0.15 (0.10–1.40) | 4.55 (0.15–9.25) | 4.40 (1.03–7.90) | 1.60 (0.20–6.60) | 1.80 (0.10–6.80) |
| Upright acid exposure time (%/24h) |  | 0.35 (0.15–2.05) | 3.35 (0.15–7.90) | 3.05 (1.00–7.48) | 2.60 (0.20–6.50) | 2.60 (0.20–6.85) |
| Supine acid exposure time (%/24h) |  | 0.00 (0.00–0.00) | 4.05 (0.00–9.23) | 0.60 (0.00–9.18) | 0.00 (0.00–5.40) | 0.20 (0.00–2.65) |
| Mean acid clearance time (min) |  | 0.80 (0.43–1.48) | 1.60 (0.90–2.00) | 2.20 (1.15–3.75) | 1.80 (1.10–2.70) | 1.35 (0.43–2.15) |
| DeMeester score | Normal <14.72 | 1.45 (1.08–5.20) | 17.85 (1.03–34.90) | 14.20 (3.10–35.00) | 4.60 (1.10–29.65) | 6.15 (1.15–22.50) |
| Total reflux episodes (n/24h) | Normal <40 | 29.50 (18.75–49.25) | 39.00 (25.25–53.75) | 39.50 (22.75–62.00) | 25.00 (21.00–54.00) | 51.00 (30.00–81.50) |
| Acid reflux episodes (n/24h) |  | 2.00 (1.00–3.75) | 6.00 (2.25–22.75) | 3.50 (2.00–12.25) | 2.00 (1.00–4.00) | 4.00 (2.00–15.50) |
| Weakly acidic reflux episodes (n/24h) |  | 20.00 (12.50–31.25) | 20.00 (12.50–28.50) | 23.50 (19.00–43.75) | 21.00 (16.00–39.00) | 34.00 (20.00–67.50) |
| MNBI (Ω) | Abnormal <2500 | 1847 (1404–2085) | 792.5 (484.5–1165) | 800.5 (332–1227) | 839 (336–1319) | 1265 (815–1538) |

**Footnote:** Values are presented as median (interquartile range, IQR). Total acid exposure time (AET) represents the percentage of time with esophageal pH <4 over 24 hours; upright and supine AET correspond to acid exposure during upright and recumbent periods, respectively. The DeMeester score is a composite measure of esophageal acid exposure, with values <14.72 considered normal. Impedance reflux episodes represent the total number of reflux events detected over 24 hours, including acid (pH <4) and weakly acidic (pH 4–7) episodes. Mean acid clearance time reflects the duration required for esophageal pH to return above 4 following a reflux event. The longest reflux episode corresponds to the maximum duration of a single reflux event. Mean nocturnal baseline impedance (MNBI) reflects esophageal mucosal integrity and/or stasis.

Figure 3 depicts pH/MII parameters according to isolated and combined esophageal and gastric motor dysfunction. Patients with isolated esophageal dysmotility demonstrated higher AET, longer clearance times, and a greater proportion of conclusive AET values. Those with isolated gastric dysmotility had increased reflux episode counts, particularly weakly acidic reflux, without consistent elevations in AET. Patients with combined esophageal and gastric dysmotility exhibited the most severe phenotype with higher AET, increased reflux burden, and prolonged clearance times. In contrast, patients with normal esophageal and gastric motility (n=2) showed pH/MII parameters within physiological ranges.

**Figure 3:**
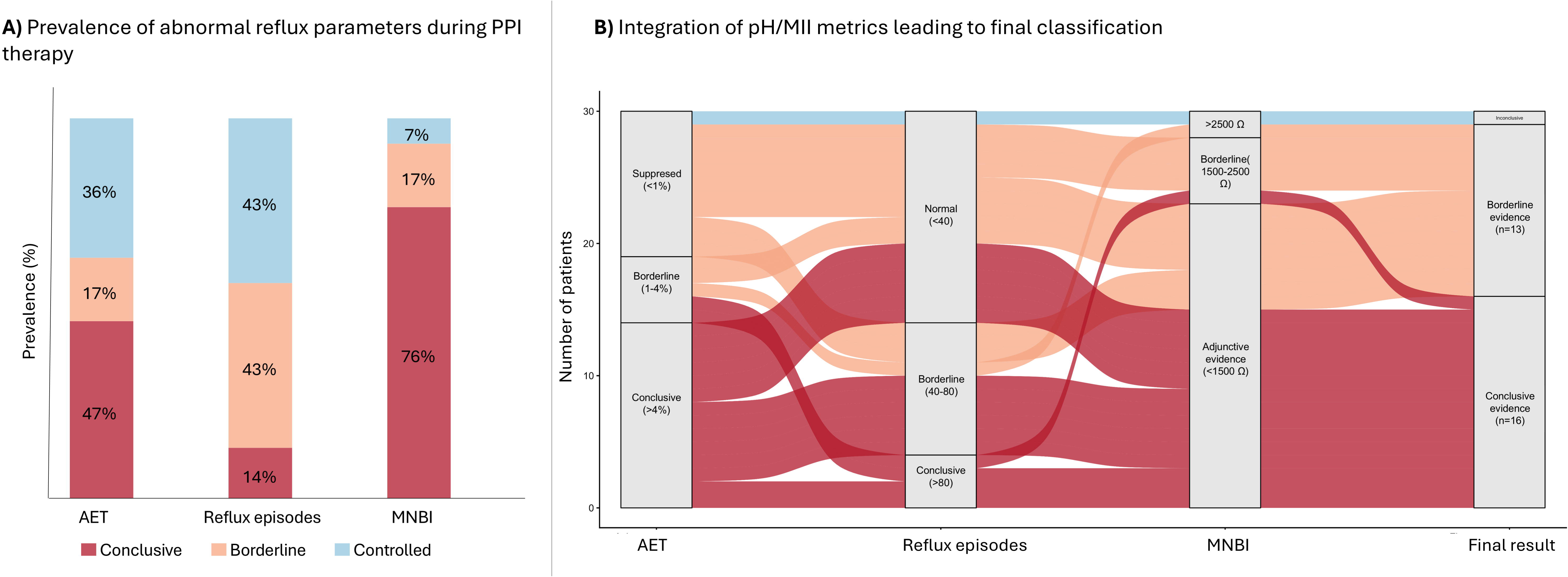
Distribution of reflux burden, clearance, and mucosal integrity across esophageal and gastric motility phenotypes. Footnote: Boxplots depict the distribution of (A) acid exposure time (AET), (B) number of reflux episodes, (C) mean acid clearance time, and (D) mean nocturnal baseline impedance (MNBI) across groups defined by esophageal and gastric motility status: both results normal, esophageal dysmotility only, gastric dysmotility only, and combined esophageal and gastric dysmotility. Individual data points are color-coded according to lyon 2.0 thresholds: AET (conclusive >4%, borderline 1–4%, suppressed <1%), reflux episodes (conclusive >80, borderline 40–80, normal <40), MNBI (low <1500 Ω, borderline 1500–2500 Ω, high >2500 Ω), and acid clearance time (<2 min vs ≥2 min). For visualization purposes, the y-axis for acid clearance time is truncated at 15 minutes, one patient (not shown) from the combined group had a Mean clearance time of 28 min.

## DISCUSSION

The main clinically relevant finding of this study is that objective evidence of ongoing GERD was present in nearly all patients with SSc, including those undergoing evaluation for lung transplantation and those without persistent reflux symptoms, despite treatment with high-dose PPI therapy. This prevalence markedly exceeds that reported in non-SSc populations and underscores that GERD in SSc frequently persists despite aggressive acid suppression. Importantly, our findings demonstrate that PPI-refractory GERD in SSc is mechanistically heterogeneous. Esophageal motor impairment, especially absent contractility, was associated with impaired acid clearance and increased cumulative acid exposure, whereas gastric dysmotility was linked to an increased reflux episode burden, including weakly acidic events that are not mitigated by PPIs. Together, these complementary yet distinct pathophysiologic patterns indicate that refractory GERD in SSc arises from coordinated dysfunction across multiple foregut compartments rather than from isolated esophageal abnormalities. This multi-factorial mechanism provides a compelling explanation for the limited effectiveness of acid suppression monotherapy in many patients with SSc.

In our cohort, most patients with apparently normal acid exposure on PPI therapy nevertheless exhibited abnormal MNBI values, consistent with prior studies reporting reduced MNBI and increased reflux burden in SSc patients undergoing pH/MII while on PPI therapy^34^. MNBI has been proposed as a stable surrogate marker of mucosal integrity and cumulative reflux exposure,^33^ suggesting persistent reflux-related mucosal impairment even when acid control appears adequate. However, interpretation of MNBI in SSc warrants caution. Low MNBI values may also reflect luminal stasis of esophageal contents, as impaired clearance can prolong mucosal contact time and reduce impedance independent of acid exposure.^35,36^. Accordingly, in SSc, reduced MNBI may represent the combined effects of reflux exposure, nocturnal stasis, and ineffective clearance rather than from acid injury alone. From a patient perspective, nocturnal regurgitation in the supine position and esophageal stasis are particularly clinically relevant, as they often drive symptom burden despite escalated acid suppression.^14,37^. Thus, although the relative contribution of each mechanism cannot be disentangled in this study, abnormal MNBI in SSc likely reflects the intersection of impaired clearance and ongoing reflux rather than acid exposure alone.

These findings have important implications for clinical practice. In the general population, current guidelines recommend confirming GERD with testing performed off PPI-therapy^19,20^. In SSc, however, PPI withdrawal is often poorly tolerated due to severe rebound symptoms and the heightened risk of aspiration-related pulmonary injury^13,38^. Given the high prevalence of objective GERD and the frequent abnormalities observed on pH/MII testing performed during ongoing PPI therapy in our cohort, on-PPI pH/MII assessment appears to be a pragmatic and informative diagnostic strategy in patient with persistent symptoms or concern for clinically significant reflux. This approach may facilitate quantification of ongoing reflux burden without precipitating symptom deterioration and may better align diagnostic evaluation with the unique physiologic vulnerabilities of patients with SSc.

Beyond diagnostic considerations, our results reinforce the need for mechanism-based management strategies. The observed associations between esophageal dysmotility and impaired acid clearance, and between gastric dysmotility and increased reflux episodes, support a multifactorial model in which dysfunction across the esophagus, EGJ, and stomach contributes to refractory disease expression. Patients with combined esophageal and gastric dysfunction demonstrated the most severe abnormalities across reflux metrics, underscoring the additive impact of multi-level foregut involvement. Notably, gastric dysmotility appears particularly clinically relevant. In addition to its association with increased reflux episode burden in the present study, prior work from our group has linked delayed gastric emptying to refractory esophagitis and adverse outcomes, including lung transplantation and mortality.^11,4^. Together, these findings raise the possibility that gastric dysmotility may may serve as both a therapeutic target and a marker of higher-risk disease in SSc.

Mechanistically, delayed gastric emptying may exacerbate reflux through increased gastric volume and enhanced transient lower esophageal sphincter relaxations^39^, while esophageal hypomotility prolongs exposure to both acid and non-acid refluxate by impairing clearance^40,41^. These observations suggest that selected patients may benefit from adjunctive strategies beyond acid suppression, including prokinetic therapy in those with documented gastric dysfunction^42^, pharmacological approaches aimed at improving EGJ barrier function (e.g., buspirone or tone-enhancing prokinetics)^43^, or carefully selected anti-reflux- interventions^44^. Such approaches must be individualized and applied cautiously, particularly given the high prevalence of severe esophageal dysmotility and the attendant risk of post-intervention dysphagia.

This study is not without limitations. Gastrointestinal motility testing and pH/MII monitoring were not contemporaneous, reflecting real-world practice but introducing potential temporal variability. Patient-reported outcome (PRO) measures were not available at the time of physiologic testing, limiting symptom-physiology correlations. The very high prevalence of objective refractory GERD also precluded meaningful comparisons with patients with well-controlled disease. Finally, the cross-sectional design of the study limits causal inference. Prospective studies integrating longitudinal symptom assessment with physiologic testing and clinical outcomes are needed to define clinically actionable thresholds and validate phenotype-guided treatment strategies.

In summary, this study provides a comprehensive physiologic characterization of PPI-refractory GERD in SSc, using integrated esophageal, EGD, and gastric assessments performed during ongoing PPI therapy. Our findings demonstrate that refractory GERD in SSc is highly prevalent and driven by distinct, quantifiable abnormalities across the upper gastrointestinal tract. Clinically, these findings support a shift toward individualized, mechanism-based management strategies that extend beyond acid suppression alone and lay the foundation for future trials targeting dominant physiologic drivers of disease.

## Data Availability

Upon reasonable request

## ETHICAL CONSIDERATIONS

All methods were performed in accordance with relevant guidelines and regulations, and the study was approved by the Hospital Vall d’Hebron Ethics Committee for Clinical Research [PR(AG)312/2021]. Patients gave written informed consent for the management of clinical data.

## ACKNOWLEDGEMENTS

Dr Hughes is supported by the National Institute for Health and Care Research (NIHR) Manchester Biomedical Research Centre (BRC) (NIHR203308).

## DATA SHARING STATEMENT

Data are available upon reasonable request.

## AUTHOR CONTRIBUTIONS

All authors meet all four ICMJE criteria. LGAG, AGdC: study conception and design, study coordination, data acquisition, statistical analysis, and manuscript drafting. FAFT: data acquisition, data analysis, and manuscript drafting. AA, CB, CM, LPF, LT, CCC: data acquisition. CPSA, AGdC, JS: data interpretation, critical revision of the manuscript, and supervision. ZM: study conception and design, data interpretation, and manuscript drafting. All authors critically revised the manuscript for important intellectual content and approved the final version.

